# Efficacy and Safety of Wharton’s Jelly-Derived Mesenchymal Stem Cells in Patients with Ischemic Cardiomyopathy: A Randomized Pilot Trial

**DOI:** 10.64898/2026.07.03.26357234

**Authors:** Luis Horacio Atehortua, Sergio Estrada-Mira, Oscar Velazquez, Juan Pablo Florez, Francisco Villegas, Santiago Torres-Alzate, Mauricio Atehortua, Oscar Villada, Juan Camilo Ortiz, Fabian Jaimes

## Abstract

**Introduction:** Wharton’s jelly-derived mesenchymal stem cells (WJ-MSCs) have emerged as a promising regenerative strategy for ischemic heart disease because of their immunomodulatory, angiogenic, and antifibrotic properties. This pilot randomized trial evaluated the safety, feasibility, and exploratory efficacy of intramyocardial WJ-MSC administration combined with an extracellular matrix (ECM) patch in patients with ischemic cardiomyopathy undergoing coronary artery bypass grafting (CABG).

**Methods:** In this randomized, controlled pilot trial, 28 patients with ischemic cardiomyopathy, left ventricular ejection fraction (LVEF) <40%, and viable myocardium on cardiac magnetic resonance imaging (MRI) were assigned to receive intramyocardial WJ-MSC injections plus an extracellular matrix (ECM) patch or a placebo patch. Patients were followed for 12 months with echocardiography, cardiac MRI, Holter monitoring, functional assessment, and quality-of-life evaluation.

**Results:** Among 44 screened patients, 28 were randomized (16 to WJ-MSC and 12 to control). At 12 months, echocardiography showed a greater improvement in LVEF in the WJ-MSC group than in the control group (8% vs. 0%, p=0.045). Myocardial fibrosis decreased by 32% in both groups. Cardiac MRI demonstrated improvement in both groups, with numerically greater gains in LVEF and larger reductions in fibrosis in the WJ-MSC arm, although between-group differences were not statistically significant. No significant between-group differences were observed in ventricular arrhythmias or serious adverse events. Two non-cardiac postoperative deaths occurred in the WJ-MSC group.

**Conclusions:** Intramyocardial WJ-MSC administration combined with an ECM patch during CABG appears feasible and safe, with signals of functional improvement. Larger, adequately powered trials are needed to confirm efficacy and long-term safety.

**Trial registration:** ClinicalTrials.gov, NCT04011059.

## Introduction

The contemporary paradigm of cardiac regeneration has shifted from a cell replacement-centered approach to a functional model prioritizing microenvironment modulation, structural support, and the paracrine effects of stem cells. Wharton’s jelly-derived mesenchymal stem cells (WJ-MSCs) have emerged as a promising source due to their immunomodulatory, angiogenic, and antifibrotic properties, mediated by a secretome rich in bioactive factors and extracellular vesicles.(1–5)

Furthermore, the use of three-dimensional biological scaffolds such as extracellular matrix (ECM) patches has shown to significantly enhance cell retention and tissue interaction, thereby potentiating therapeutic effects. In experimental models, the synergistic combination of WJ-MSCs with these structural patches has demonstrated positive impacts on tissue organization, perfusion, and overall contractility of the injured myocardium.(6,7)

The concept of ‘functional regeneration’ has thus been consolidated, recognizing that restoring cardiac function goes beyond integrating new cells. It involves a multifactorial intervention that improves contractility, ventricular architecture, and the cellular microenvironment. This study, by simultaneously integrating WJ-MSCs and a bioactive epicardial patch during surgery, represents progress toward a clinically viable and comprehensive regenerative therapy model.(3,6,8)

Ischemic heart disease remains one of the leading causes of morbidity and mortality worldwide. Although therapeutic advances in the acute phase of myocardial infarction have reduced immediate lethality, a high percentage of patients survive with irreversible myocardial damage, leading to the progressive development of heart failure. This condition represents not only a significant clinical burden but also an increasing economic and social challenge.(4,9)

For decades, the heart was believed to be a post-mitotic organ incapable of regeneration. However, recent studies have shown that adult cardiomyocytes possess a limited but existent capacity for proliferation. This understanding has shifted research efforts toward strategies aimed not only at halting functional deterioration but also at promoting effective myocardial tissue regeneration.(10,11)

Despite numerous clinical trials with cell therapies, the observed benefits have been modest. It is now known that these effects derive primarily from paracrine mechanisms rather than from the structural integration of injected cells. This reinforces the need to select cells with robust secretory, immunomodulatory, and angiogenic profiles—such as Wharton’s jelly-derived mesenchymal stem cells.(1,12)

In parallel, tissue engineering and the use of three-dimensional ECM scaffolds have shown notable improvements in the retention and efficacy of cell-based therapies. At the same time, it has become clear that the post-ischemic microenvironment—characterized by fibrosis, inflammation, and poor perfusion—significantly limits the success of these strategies. Thus, a combined approach is essential: functional cells + structural scaffolding + microenvironment modulation.(4,5,13)

There remains a knowledge gap in Latin America regarding advanced therapies using allogeneic cells such as WJ-MSCs, particularly their application in open-heart surgery and their combined effect with ECM patches. This pilot randomized clinical trial aims to evaluate the functional impact of this strategy in patients with severe ischemic heart disease and contribute to the advancement of regenerative medicine in our region.

In the past two decades, the field of cardiac regeneration has made notable progress, driven by five key lessons that redefine the contemporary therapeutic approach: (1) adult cardiomyocytes have a limited but real capacity to re-enter the cell cycle and proliferate under certain molecular stimuli; (2) the postnatal inflammatory and metabolic microenvironment, including reactive oxygen species (ROS) production, plays a determining role in halting endogenous myocardial regeneration, thereby interfering with the regeneration that is still possible in the neonatal stage; (3) the impact of stem cells is mainly paracrine, mediated by exosomes and soluble factors rather than structural integration; (4) direct reprogramming and epigenetic editing open new pathways to convert resident somatic cells into functional cardiomyocytes; and (5) the use of three-dimensional structural supports and tissue environment modulation are essential conditions for the success of any regenerative strategy in damaged myocardial tissue. These lessons have reshaped clinical goals, prioritizing functional heart regeneration over traditional cell replacement.(4,5,9)

This knowledge has motivated strategies such as direct reprogramming of fibroblasts into cardiomyocytes, and the use of genetic factors, modified RNAs, and extracellular vesicles as emerging regenerative tools. Likewise, the use of bioactive cardiac patches seeded with pluripotent stem cell-derived cardiomyocytes (PSC-CMs) has shown promising results, although challenges remain in cell maturation and electrical safety, as evidenced by the appearance of ventricular arrhythmias in animal models and preclinical studies. The electrical immaturity of PSC-CMs and their tendency to generate ectopic foci have led to the development of new technologies for ‘functional pre-training’ and co-culture with supporting cells, as well as the use of bioactive matrices that promote better electrical and mechanical integration with host tissue.(1,4,9)

Despite these challenges, the combination of stem cells, smart biomaterials, and controlled biochemical signals appears to be the most promising avenue for achieving sustained structural and functional regeneration of injured myocardium.

This pilot clinical trial fits within this context of advancements and provides initial evidence on a combined strategy using an epicardial patch with WJ-MSCs to promote functional myocardial regeneration in patients with ischemic cardiomyopathy, simultaneously addressing the structural, immunological, and bioelectrical limitations of the damaged tissue.(14)

## Methods

### Study Design

This was a randomized, controlled, quadruple-masked, proof-of-concept pilot clinical trial with a parallel two-group design. The manuscript follows CONSORT 2025 guidance for randomized trials and incorporates pilot/feasibility reporting considerations. The intervention was evaluated as an adjunct to coronary artery bypass grafting (CABG) in patients with ischemic cardiomyopathy.(15)

Patient and public involvement: Patients and members of the public were not formally involved in the design, conduct, reporting, or dissemination planning of this pilot clinical trial.

### Trial registration, recruitment, and follow-up period

The trial was prospectively registered at ClinicalTrials.gov (NCT04011059). The ClinicalTrials.gov PRS release receipt is dated July 5, 2019, when the record status was “Not yet recruiting”. The actual study start recorded in ClinicalTrials.gov was November 23, 2019; therefore, no participant was enrolled before public trial registration.

According to the final study assignment/procedure records, participant treatment/procedure data collection for the randomized study began on November 29, 2019 and ended on October 28, 2022. Participants underwent protocol-specified follow-up at 3, 6, 9, and 12 months after surgery; the final 12-month follow-up window therefore extended through October 28, 2023. The authors confirm that all ongoing and related trials for this intervention are registered.

### Ethics and informed consent

The study was approved before participant enrollment by the Comité de Ética de la Investigación, Fundación Hospitalaria San Vicente de Paúl, Medellín, Colombia, as recorded in Minutes No. 31-2018, approved on November 9, 2018. The committee subsequently approved the expansion of protocol execution to Fundación Hospital San Vicente de Paúl-Rionegro as recorded in Minutes No. 33-2019, approved at the committee meeting of November 15, 2019, with notification dated November 27, 2019. All participants provided written informed consent before inclusion, including consent to participate and to publish aggregated study results. All procedures were conducted in accordance with the Declaration of Helsinki and applicable Colombian regulations.

### Protocol deviations

The registered protocol anticipated enrollment of 40 participants, with 20 participants in each treatment arm over an approximately 36-month recruitment period. Because of the limited eligible population, operational feasibility constraints related to cell processing and intraoperative administration, and the completion of the funded study period, 28 participants were ultimately randomized (16 to WJ-MSC and 12 to control). This deviation affected only the achieved sample size and allocation balance. It did not change the randomized design, eligibility criteria, intervention components, planned outcome definitions, or the protocol-specified follow-up schedule. Accordingly, all efficacy analyses were considered exploratory and hypothesis-generating.

The primary objectives were to evaluate changes from baseline in left ventricular structure and function and to assess ventricular arrhythmic events during postoperative follow-up at 3, 6, 9, and 12 months. Specifically, we aimed to:

- Assess changes from baseline in left ventricular ejection fraction (LVEF), as measured by transthoracic echocardiography and cardiac magnetic resonance imaging (CMR);
- Assess changes from baseline in left ventricular end-diastolic and end-systolic volumes, as measured by echocardiography and CMR;
- Evaluate changes in myocardial viability, defined by segmental wall motion improvement and affected wall thickness on CMR; and
- Determine the incidence of ventricular arrhythmias, defined as non-sustained ventricular tachycardia and high- or low-grade ventricular premature beats.

### Secondary objectives

The secondary objectives were to evaluate changes in functional capacity, quality of life, myocardial fibrosis, exercise tolerance, and mortality during follow-up. Specifically, we aimed to:

- Assess recovery of functional status according to the New York Heart Association (NYHA) functional classification at 3, 6, 9, and 12 months;
- Evaluate changes from baseline in the Minnesota Living with Heart Failure Questionnaire (MLHFQ)(16) score at 3, 6, 9, and 12 months;
- Assess changes in late gadolinium enhancement on CMR, expressed as the percentage of wall thickness affected in each segment, at 3, 6, 9, and 12 months;
- Evaluate changes from baseline in the distance covered during the 6-minute walk test at 3, 6, 9, and 12 months;
- Determine cardiovascular mortality at 3 and 12 months;
- Determine all-cause mortality at 3 and 12 months.

Harms and safety assessment: Harms and safety outcomes included perioperative serious adverse events, all-cause and cardiovascular mortality, and ventricular arrhythmic events assessed by protocol-specified Holter monitoring. Events were captured from clinical follow-up, medical records, mortality surveillance, and scheduled rhythm assessment during the 12-month follow-up period.

Participants: Patients aged 30 to 75 years with ischemic cardiomyopathy eligible for coronary revascularization. Included patients had LVEF <40% with segmental motion abnormalities and viable myocardium identified by cardiac MRI. Exclusion criteria included severe arrhythmias, active cancer, or multi-organ failure.

Interventions: The experimental group received 8 x10^7^ intramyocardial WJ-MSC injections and an extracellular matrix (ECM) patch seeded with 1 x 10^7^ WJ-MSCs. The control group received culture medium injection and a patch without cells. Injections were image-guided and targeted ischemic regions identified by MRI.

Concomitant care: Both groups underwent CABG and received standard perioperative, postoperative, and heart-failure care according to institutional clinical practice and treating physician judgment.

Sample Size: The registered protocol anticipated 40 participants (20 per arm). The achieved sample size was 28 participants (16 WJ-MSC and 12 control), reflecting the pilot, feasibility-oriented nature of the trial and the limited number of eligible patients during the study period. No formal a priori sample size calculation was performed for efficacy outcomes. Therefore, the trial was not powered to detect definitive between-group differences, and efficacy analyses were considered exploratory.

Randomization and allocation concealment: Allocation was determined using a random number generator (Ralloc, Stata) with randomly permuted blocks of 2, 4, and 6. Treatment allocation was known only to the tissue bank/cell-processing team responsible for preparing and delivering the syringes and epicardial patches. Participants, clinical care providers, investigators, and outcome assessors were masked to group assignment.

Statistical Analysis: Categorical variables were expressed as absolute numbers and percentages. Numerical variables were reported as medians with interquartile ranges. Between-group differences were tested using the Mann-Whitney U test or Student’s t-test for numerical variables based on distribution assumptions, and Pearson’s chi-squared or Fisher’s exact test for categorical variables. Wilcoxon test was used for intragroup repeated measures. A p-value < 0.05 was considered statistically significant. All analyses were performed with STATA. No formal interim efficacy analyses, stopping guidelines, subgroup analyses, or sensitivity analyses were prespecified or performed. Missing follow-up outcome data were not imputed; efficacy analyses used available outcome data at each time point, whereas safety analyses included all randomized participants whenever data were available.

Ethical Approvals: The study was approved by the Comité de Ética de la Investigación, Fundación Hospitalaria San Vicente de Paúl, Medellín, Colombia, as recorded in Minutes No. 31-2018, approved on November 9, 2018. The committee subsequently approved the expansion of protocol execution to Fundación Hospital San Vicente de Paúl-Rionegro as recorded in Minutes No. 33-2019, approved at the committee meeting of November 15, 2019, with notification dated November 27, 2019. The trial was registered at ClinicalTrials.gov (NCT04011059). Written informed consent was obtained from all participants before enrollment.

## Results

A total of 44 patients scheduled for coronary artery bypass grafting (CABG) were evaluated. Of these, 14 were excluded for not meeting eligibility criteria and 2 were not randomized. Ultimately, 28 patients were enrolled and randomized in the study: 16 in the WJ-MSC group and 12 in the placebo/control group (Fig 1).

**Fig 1.**
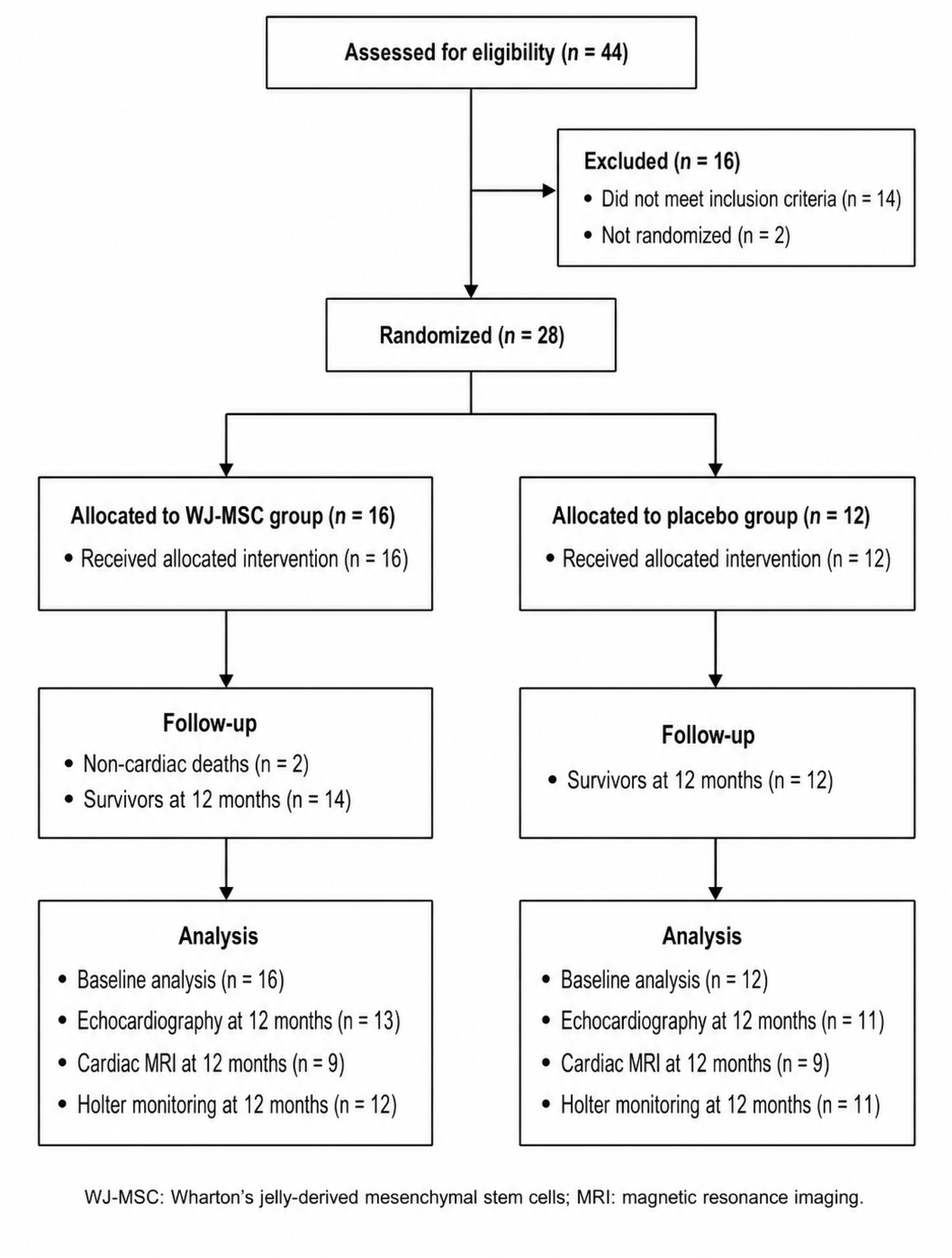
CONSORT flow diagram. Flow of participants through eligibility assessment, randomization, allocation, follow-up, and analysis. CABG: coronary artery bypass grafting; CMR: cardiac magnetic resonance imaging; ECM: extracellular matrix; WJ-MSC: Wharton’s jelly-derived mesenchymal stromal/stem cell.

**Table 1.** shows the demographic characteristics and clinical history of the study groups.

| Variable | WJ-MSC (n=16) | Placebo (n=12) |
| --- | --- | --- |
| Male gender (%) | 75 | 91.7 |
| Age (years), Median (IQR) | 65 (60.2–68.7) | 61.5 (59.2–72.2) |
| Weight (kg), Median (IQR) | 65.5 (62–73.7) | 67 (64.2–78.7) |
| Height (cm), Median (IQR) | 165.5 (160–170) | 166.6 (161–170) |
| Diabetes (%) | 37.5 | 41.7 |
| COPD (%) | 12.5 | 0 |
| Hypertension (%) | 81.3 | 50 |
| Renal insufficiency (%) | 18.8 | 8.3 |
| Obesity (%) | 12.5 | 8.3 |
| Dyslipidemia (%) | 62.5 | 33.3 |
| Smoker (%) | 25 | 41.7 |
| History of coronary disease (%) | 25 | 0 |
| Three-vessel disease (%) | 81.3 | 91.7 |
| CABG surgery (%) | 93.8 | 91.7 |
| Hemoglobin (g/dL), Median (IQR) | 11.95 (11.3–14.6) | 12.25 (11–13.2) |
| Hematocrit (%), Median (IQR) | 35.9 (33.4–42.3) | 34.35 (31.4–37.3) |
| Creatinine (mg/dL), Median (IQR) | 1.03 (0.84–1.09) | 0.865 (0.73–1.09) |

Me: Median; IQR: Interquartile Range; COPD: Chronic Obstructive Pulmonary Disease; CABG: Coronary Artery Bypass Grafting.

### Primary endpoint

At 12-month follow-up, echocardiographic evaluation showed significant improvement in the median left ventricular ejection fraction (LVEF) in the WJ-MSC group (from 36% to 44%, p=0.045), while the placebo group showed no significant change (38% baseline and follow-up). Additionally, improvements were observed in end-diastolic volume, end-systolic volume, and global longitudinal strain in the WJ-MSC group, indicating enhanced systolic function. (Table 2)

**Table 2.**
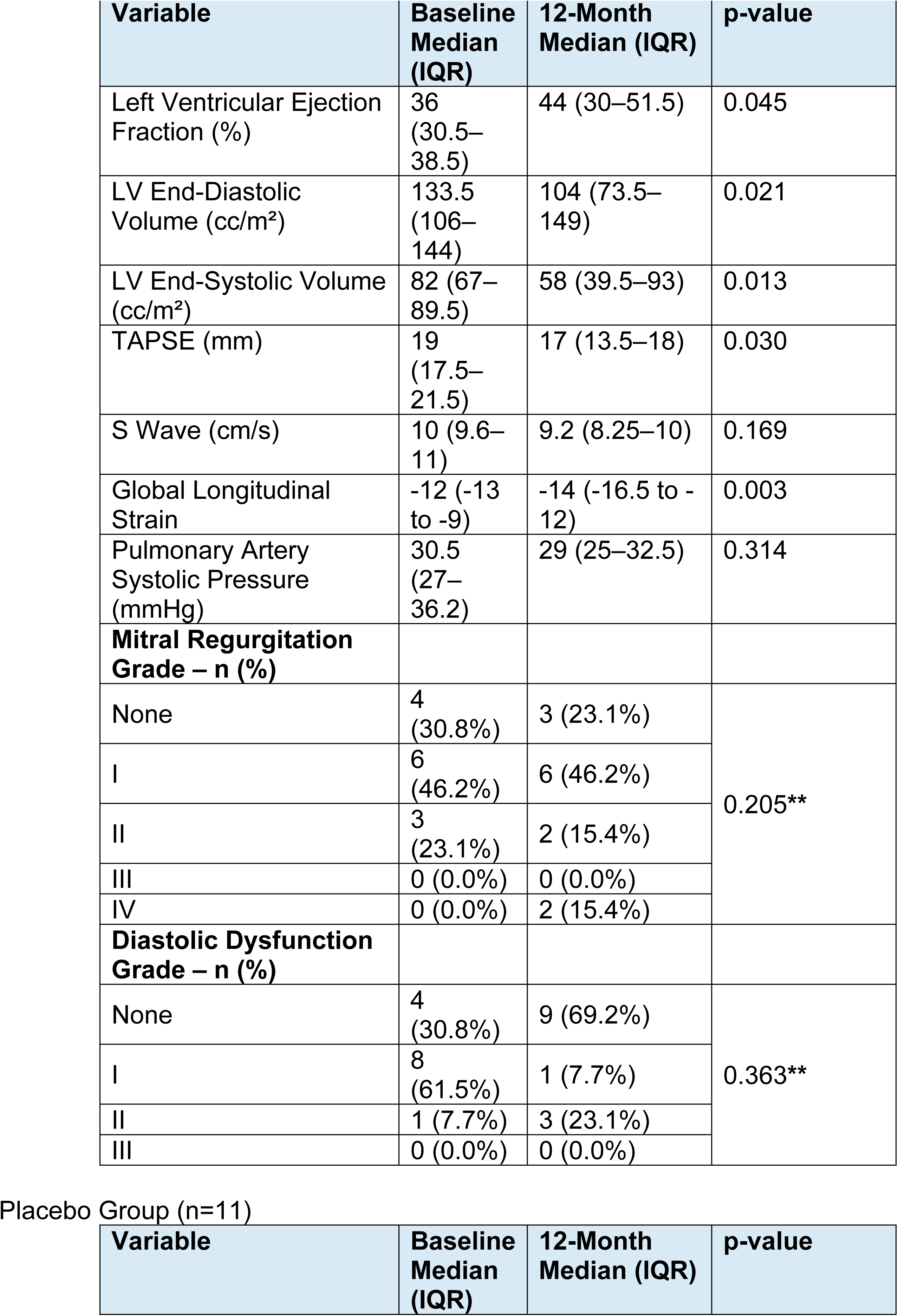
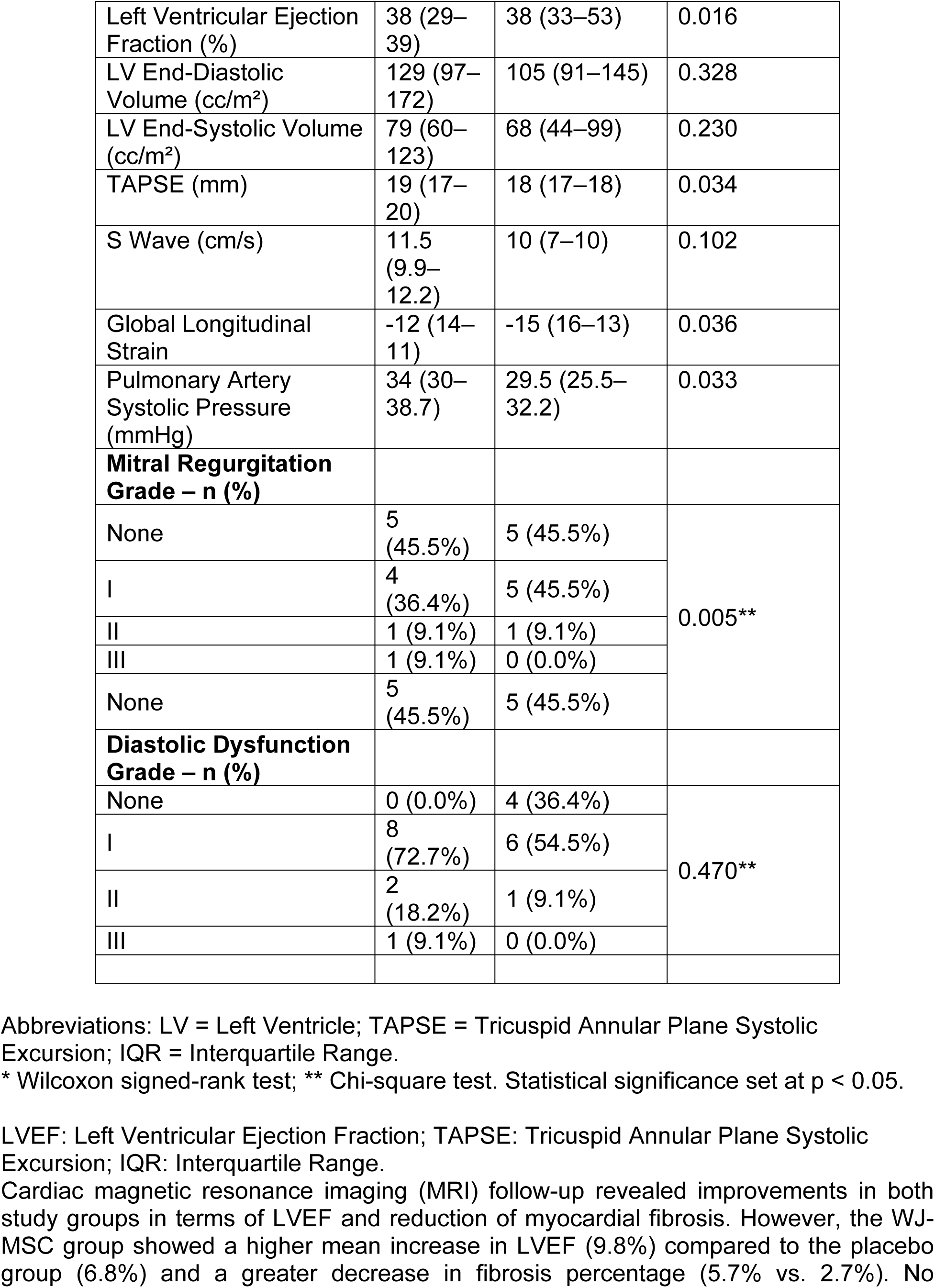
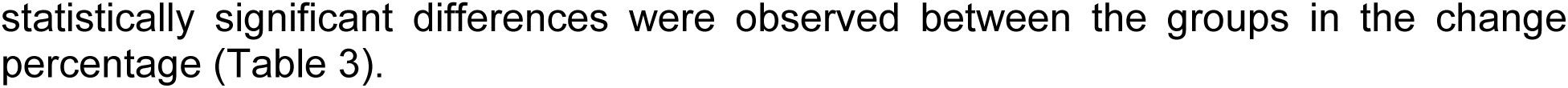
Echocardiographic Evaluation: Baseline and 12 Months Mesenchymal Stem Cells Group (n=13).

**Table 3.** Changes in Cardiac MRI Parameters from Baseline to 12 Months by Group.

| **Variable** | **WJ-MSC**<br>**n=9**<br>**Mean (SD)** | **Placebo**<br>**n=9**<br>**Mean (SD)** | **p-value\*\*** |
| --- | --- | --- | --- |
| Left Ventricular Ejection Fraction (LVEF, %) |  |  |  |
| Baseline | 33.4 (4.4) | 34.6 (3.7) |  |
| Month 12 | 43.2 (9.8) | 41.4 (8.5) |  |
| % Change (Baseline vs. 12 months) | 9.8 (6.7) | 6.8 (7.6) | 0.391 |
| Global Wall Motion Index |  |  |  |
| Baseline | 1.83 (0.5) | 1.68 (0.2) |  |
| Month 12 | 1.63 (0.3) | 1.62 (0.3) |  |
| Mean Difference* | -0.2 (0.7) | -0.05 (0.2) | 0.602 |
| Fibrosis (%) |  |  |  |
| Baseline | 23 (12.5) | 26 (15.6) |  |
| Month 12 | 17.3 (9.2) | 23.3 (15.3) |  |
| % Change* | -5.7 (11.7) | -2.7 (3.3) | 0.482 |
| LV Systolic Volume (cc/m<sup>2</sup>) |  |  |  |
| Baseline | 107.3 (38.4) | 104.1 (35) |  |
| Month 12 | 79.4 (29.8) | 90.2 (33.9) |  |
| Mean Difference* | -27.8 (30) | -14 (26.7) | 0.312 |
| LV Diastolic Volume (cc/m<sup>2</sup>) |  |  |  |
| Baseline | 105.3 (30) | 109 (23) |  |
| Month 12 | 140.3 (38) | 152.4 (47.3) |  |
| Mean Difference* | 35 (27) | 43 (35) | 0.331 |

LVEF: Left Ventricular Ejection Fraction; SD: Standard Deviation. Negative values indicate a decrease from baseline; positive values indicate an increase.

**Table 4.** Holter Monitoring Results Pre-Surgery and at 12-Month Follow-up.

| **Variable** | **WJ-MSC** | **Placebo** | **p-value** |
| --- | --- | --- | --- |
| Cardiac Arrhythmias - Preoperative | 9/12 (75%) | 7/9 (77.8%) |  |
| Ventricular Extrasystoles - Preoperative | 7/12 (58.3%) | 8/9 (88.9%) |  |
| High-Grade Ventricular Arrhythmia - Preoperative | 3/12 (25%) | 6/9 (66.7%) |  |
| Non-Sustained Ventricular Tachycardia (Monomorphic) - Preop | 2/12 (16.7%) | 2/9 (22.2%) |  |
| Sustained Ventricular Tachycardia - Preop | 0/12 | 0/9 |  |
| Cardiac Arrhythmias - 12 Months | 10/12 (83.3%) | 8/11 (72.7%) | 0.64 |
| Ventricular Extrasystoles - 12 Months | 9/12 (75%) | 8/11 (72.7%) | 0.901 |
| High-Grade Ventricular Arrhythmia - 12 Months | 4/12 (33.3%) | 8/11 (72.7%) | 0.059 |
| Non-Sustained VT (Polymorphic) - 12 Months | 2/12 (16.7%) | 0/11 | 0.318 |
| Non-Sustained VT (Monomorphic) - 12 Months | 1/12 (8.3%) | 2/11 (18.2%) |  |
| Sustained Ventricular Tachycardia - 12 Months | 0 | 0 |  |

VT: Ventricular Tachycardia.

### Secondary endpoints

When evaluating functional class according to the New York Heart Association (NYHA) classification, a progressive improvement in functional status was observed over time in both the WJ-MSC and placebo groups (see Supplementary Material, Table S1).

The results of the 6-Minute Walk Test reflect the distance walked in meters at 3, 6, 9, and 12 months post-intervention, with greater distances recorded at all follow-up points in the WJ-MSC group, particularly at the 12-month mark. However, these differences did not reach statistical significance (Table 5).

**Table 5.**
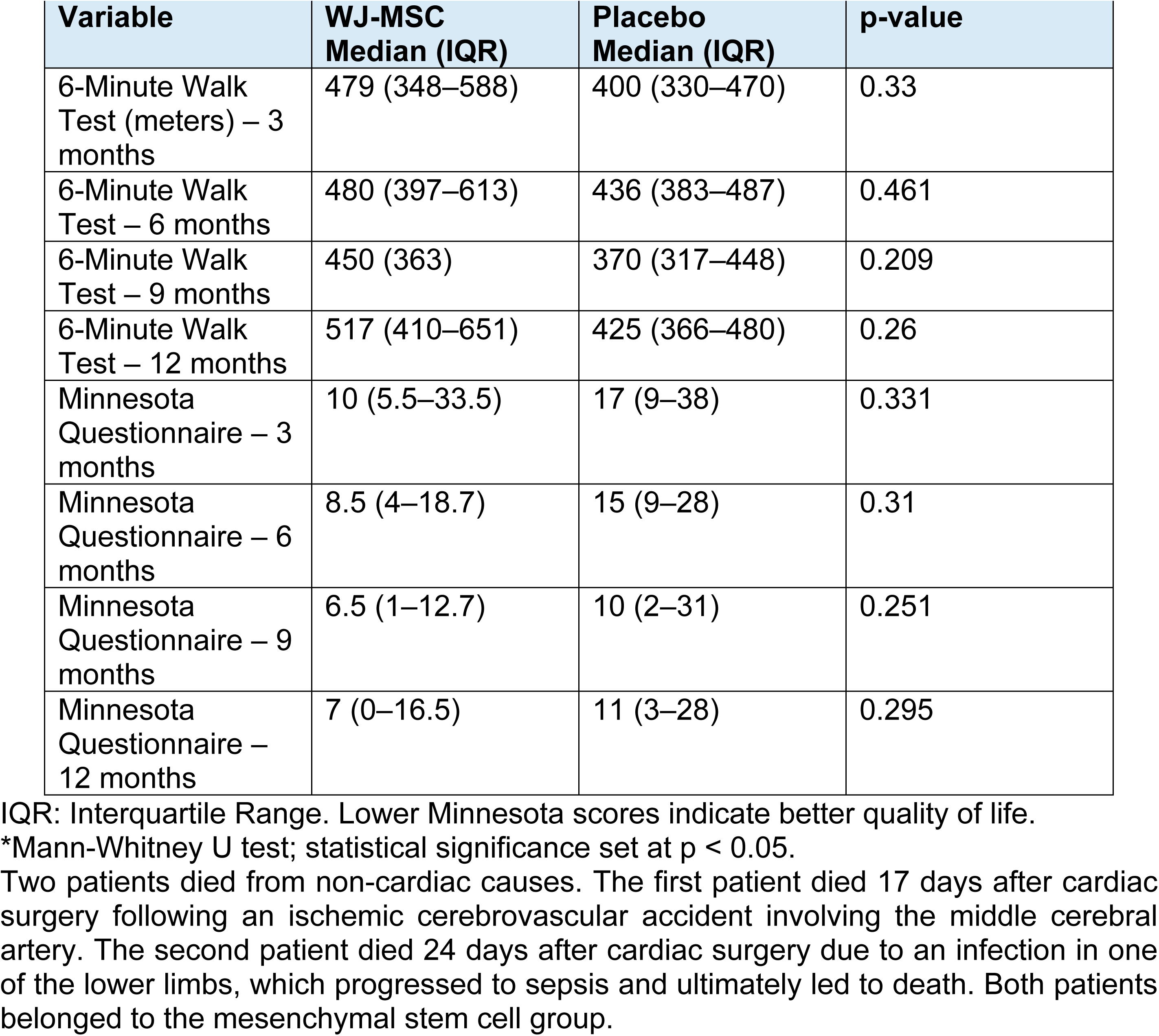
Six-Minute Walk Test and Minnesota Questionnaire Scores.

The Minnesota Living with Heart Failure Questionnaire showed lower scores at all follow-up visits in the WJ-MSC group compared to the placebo group (Table 5).

## Discussion

In addition to improvements in ejection fraction, the results suggest a favorable impact on ventricular remodeling and myocardial viability. The WJ-MSC group demonstrated a significant reduction in left ventricular end-systolic and end-diastolic volumes as assessed by echocardiography, with corresponding results in MRI, supporting both structural and functional improvement of the ventricle.

Furthermore, the global motility index analysis by MRI suggests segmental contractility improvement in the WJ-MSC group, which may indicate better recovery of ischemic segments. This could be mediated by cellular repair mechanisms, fibrosis reduction, or improved local perfusion.

The findings support that WJ-MSC administration is safe and may contribute to ventricular function recovery following coronary artery bypass grafting. Although the LVEF increase was more notable in the treatment group, the limited sample size prevents definitive conclusions.

Specifically, Holter monitoring at 12 months showed a lower incidence of high-grade ventricular arrhythmias in the WJ-MSC group (33.3%) compared to the placebo group (72.7%), with a trend toward statistical significance (p = 0.059). Likewise, the frequency of non-sustained ventricular tachycardia, whether monomorphic or polymorphic, was similar or lower in the treatment group. This reduction in complex arrhythmic events during long-term follow-up reinforces the electrical safety profile of WJ-MSC treatment, suggesting not only safety but potentially a stabilizing effect on ventricular conduction. This is particularly relevant in patients with severe systolic dysfunction, for whom arrhythmias are a major risk for adverse events.

As a potential clinical implication, the lower frequency of serious arrhythmias in the WJ-MSC group may reflect an additional benefit of this therapy, possibly reducing the risk of future cardiac complications and improving long-term outcomes.

These findings require further exploration to fully understand the mechanisms by which mesenchymal cells may influence cardiac electrophysiology and whether these effects translate into substantial clinical benefits.

Regarding NYHA functional class, a progressive improvement in functional status was observed over time in both the WJ-MSC and placebo groups. By month 12, the majority of patients were in class I, and no patients remained in class III, indicating overall improvement in both arms.

Reduction in Minnesota questionnaire scores in the WJ-MSC group indicates improvement in heart failure-related quality of life, with statistical significance observed in within-group analysis (p = 0.043). Although improvements were seen in both groups, the WJ-MSC group showed a trend toward greater benefit, especially in physical and emotional domains.

In addition to intramyocardial delivery of WJ-MSCs, a distinctive feature of this intervention was the concurrent use of an epicardial extracellular matrix (ECM) patch seeded with WJ-MSCs. This may have significantly contributed to the observed outcomes in the experimental group. ECM patches applied to the epicardial surface provide a three-dimensional bioactive microenvironment that enhances cell retention and survival, promotes differentiation and paracrine signaling, and improves integration with underlying myocardium.

Several experimental studies have shown that epicardial ECM patches combined with MSCs can induce angiogenesis, reduce fibrosis, and modulate inflammation, favoring structural and functional recovery of infarcted myocardium. The epicardial approach also facilitates sustained delivery of bioactive factors, acting as an active regenerative platform, unlike isolated injection techniques. Preclinical models have demonstrated promising results in improving ejection fraction and reducing ventricular remodeling.

In this context, the combination of intramyocardial WJ-MSC injection with epicardial patch application may have acted synergistically, amplifying the regenerative effect and contributing to the improved functional outcomes seen in the intervention group. This combined strategy may represent an advanced therapeutic approach to enhance myocardial regeneration in ischemic heart failure.

## Conclusions

Intramyocardial administration of Wharton’s jelly-derived mesenchymal stem cells (WJ-MSCs), combined with an epicardial extracellular matrix (ECM) patch during coronary artery bypass grafting (CABG), is both feasible and safe. This strategy demonstrated a favorable trend in functional recovery of the left ventricle, as evidenced by significant improvements in ejection fraction, systolic and diastolic volumes, and global longitudinal strain.

Additionally, a reduction in the incidence of high-grade ventricular arrhythmias was observed in the treated group, suggesting a favorable electrical safety profile. Quality of life, as measured by the validated Minnesota questionnaire, also improved steadily throughout follow-up.

These findings support the therapeutic potential of WJ-MSCs combined with epicardial support for functional myocardial regeneration. However, larger and statistically powered clinical trials are needed to confirm the efficacy and long-term safety of this therapeutic approach in patients with ischemic heart failure.

## Ethics declarations

### Ethics approval and consent to participate

All procedures performed in studies involving human participants were conducted in accordance with the ethical standards of the Comité de Ética de la Investigación, Fundación Hospitalaria San Vicente de Paúl, Medellín, Colombia, and with the Declaration of Helsinki. Ethics approval was granted as recorded in Minutes No. 31-2018, approved on November 9, 2018; the expansion to Fundación Hospital San Vicente de Paúl-Rionegro was approved as recorded in Minutes No. 33-2019, approved at the committee meeting of November 15, 2019, with notification dated November 27, 2019. Written informed consent to participate and to publish aggregated results was obtained from all individual participants before enrollment.

### Funding

This work was supported by Ministerio de Ciencia, Tecnología e Innovación de la República de Colombia (formerly Departamento Administrativo de Ciencia, Tecnología e Innovación [COLCIENCIAS]), Award Number 430780763747, Contract 814-2018. The funder had no role in study design, data collection and analysis, decision to publish, or preparation of the manuscript.

### Author Contributions

Conceptualization: Luis Horacio Atehortúa, Sergio Estrada Mira, Francisco Villegas, Santiago Torres and Fabián Alberto Jaimes. Laboratory analysis: Luis Horacio Atehortúa, Sergio Estrada Mira, Oscar Villada and Fabián Alberto Jaimes; Patient recruitment and clinical follow-up: Luis Horacio Atehortúa, Oscar Alberto Velázquez, Juan Pablo Flórez, Francisco Villegas, Mauricio Atehortúa and Juan Camilo Ortiz; Formal analysis and manuscript drafting: Luis Horacio Atehortúa, Sergio Estrada Mira, Oscar Villada, Santiago Torres and Fabián Alberto Jaimes. All authors reviewed and approved the final manuscript.

### Conflict of interest

All authors declare no conflicts of interest.

### Data Availability

All relevant deidentified data underlying the findings of this study are included within the manuscript and its Supporting Information files. The minimal anonymized dataset used for the analyses is provided as S3 Dataset.

## Supporting information

S1 CONSORT Checklist. Completed CONSORT 2025 checklist for this randomized pilot trial.

S2 Protocol. Trial protocol for the randomized proof-of-concept study.

S3 Dataset. Minimal deidentified dataset underlying the analyses.

## Data Availability

All relevant deidentified data underlying the findings of this study are included within the manuscript and its Supporting Information files. The minimal anonymized dataset used for the analyses is provided as S1 Dataset.

https://clinicaltrials.gov/study/NCT04011059

## Supplementary Material

**Table S1.** NYHA Functional Class by Group and Timepoint.

| NYHA | WJ-MSC |  | Placebo |  | p-value* |
| --- | --- | --- | --- | --- | --- |
|  | n | % | n | % |  |
| Baseline , n=28 | n=16 |  | n=12 |  | 0,074 |
| I | 12 | 75 | 6 | 50 |  |
| II | 1 | 6,3 | 5 | 41,7 |  |
| III | 3 | 18,7 | 1 | 8,3 |  |
| 3 months,, n=23 | n=11 |  | n=12 |  | 0,613 |
| I | 9 | 81,8 | 9 | 75 |  |
| II | 2 | 18,2 | 2 | 16,7 |  |
| III | 0 | 0 | 1 | 8,3 |  |
| 6 months,, n=20 | n=8 |  | n=12 |  | 0,477 |
| I | 8 | 100 | 10 | 83,4 |  |
| II | 0 | 0 | 1 | 8,3 |  |
| III | 0 | 0 | 1 | 8,3 |  |
| 9 months,, n=25 | n=14 |  | n=11 |  | 0,404 |
| I | 11 | 78,6 | 10 | 90,9 |  |
| II | 3 | 21,4 | 1 | 9,1 |  |
| III | 0 | 0 | 0 | 0 |  |
| 12 months,, n=24 | n=13 |  | n=11 |  | 0,482 |
| I | 11 | 84,6 | 11 | 100 |  |
| II | 2 | 15,4 | 0 | 0 |  |
| III | 0 | 0 | 0 | 0 |  |
| NYHA: New York Heart Association; *Fisher's exact test, p < 0.05 considered significant. |  |  |  |  |  |

**Table S2.**
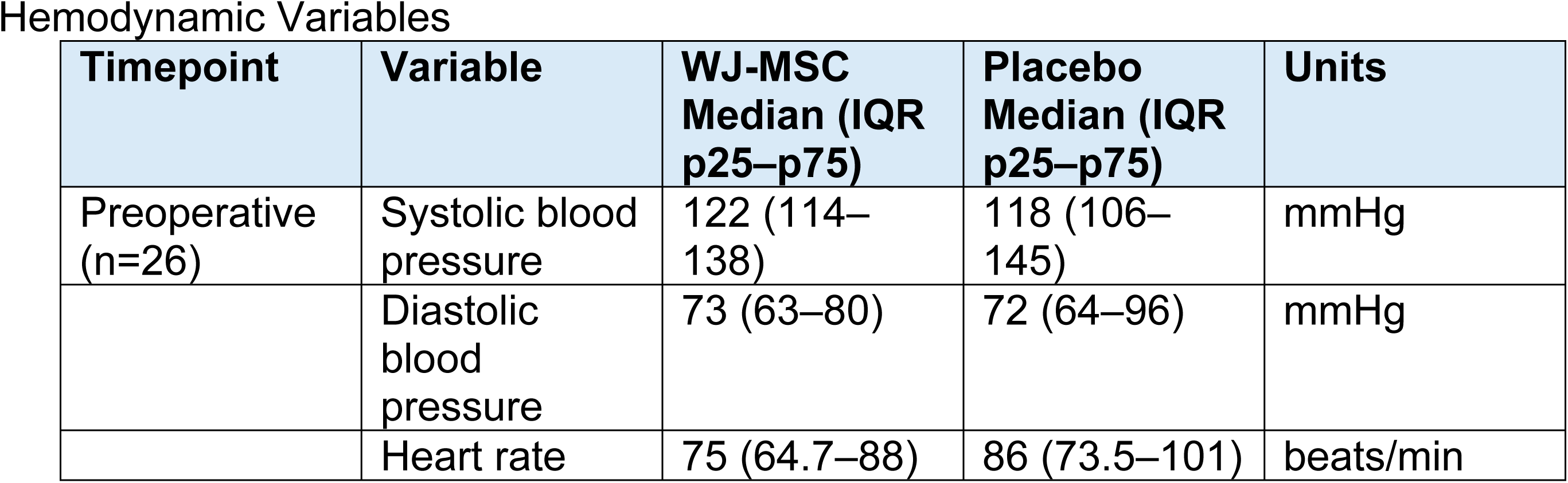
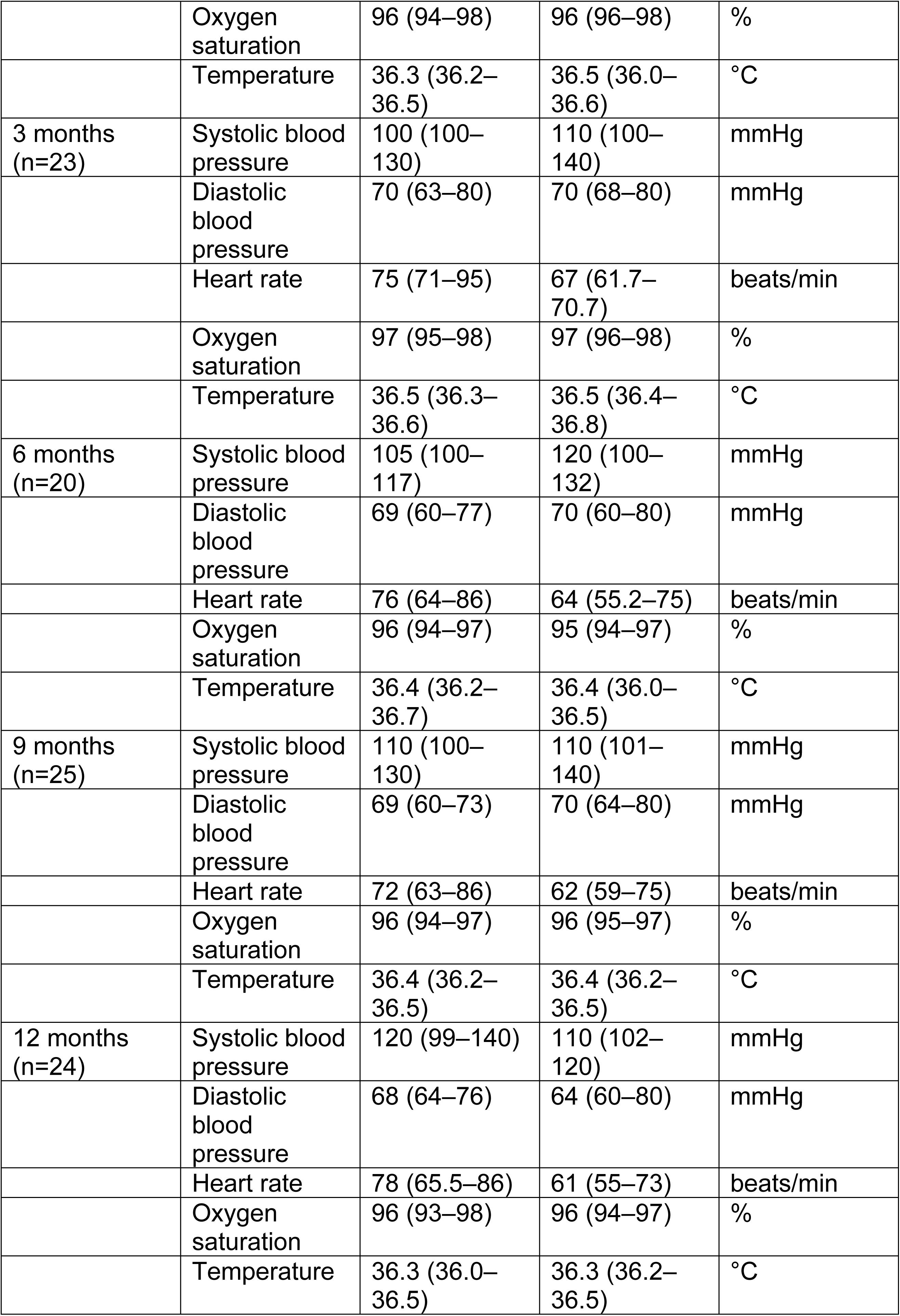
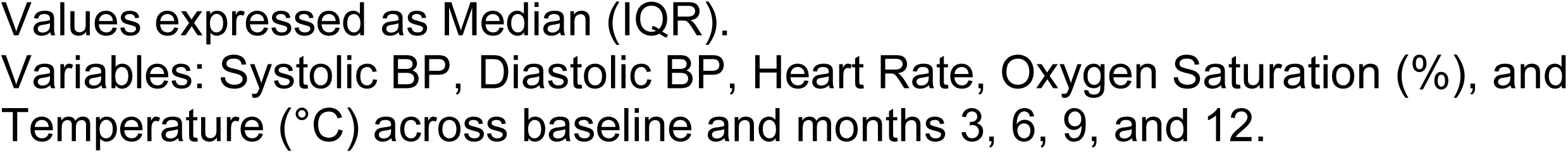
Vital Signs by Month and Treatment Group Hemodynamic Variables.

## Notes

### Competing Interest Statement

The authors have declared no competing interest.

### Clinical Trial

https://clinicaltrials.gov/study/NCT04011059

### Clinical Protocols

https://clinicaltrials.gov/study/NCT04011059

### Funding Statement

Yes

### Author Declarations

The research described in this manuscript was reviewed and approved by the Ethics Committee of Hospital San Vicente Fundación, Medellín, Colombia, under Act 14/2018, approved on 11 May 2018. The study involved human participants, and written informed consent was obtained from all participants before enrollment.

